# Cross-trial prediction of treatment response to transcranial direct current stimulation in patients with major depressive disorder

**DOI:** 10.1101/2024.09.29.24314556

**Authors:** Gerrit Burkhardt, Stephan Goerigk, Lucia Bulubas, Esther Dechantsreiter, Daniel Keeser, Ulrike Vogelmann, Katharina von Wartensleben, Johannes Wolf, Christian Plewnia, Andreas Fallgatter, Berthold Langguth, Claus Normann, Lukas Frase, Peter Zwanzger, Thomas Kammer, Carlos Schönfeldt-Lecuona, Daniel Kamp, Malek Bajbouj, Nikolaos Koutsouleris, Andre R Brunoni, Frank Padberg

**Affiliations:** Department of Psychiatry and Psychotherapy, LMU University Hospital, Munich, Germany; German Center for Mental Health (DZPG), Site Munich-Augsburg, Germany; Charlotte Fresenius Hochschule, University of Psychology, Munich, Germany; Department of Psychiatry and Psychotherapy, University Hospital, Technical University of Munich (TUM), Munich, Germany; Tübingen Center for Mental Health, Department of Psychiatry and Psychotherapy, University of Tübingen, Tübingen, Germany; German Center for Mental Health (DZPG), Site Tübingen, Germany; Department of Psychiatry and Psychotherapy, University of Regensburg, Regensburg, Germany; Department of Psychiatry and Psychotherapy, Medical Center, University of Freiburg, Freiburg, Germany; Center for Basics in Neuromodulation (NeuroModulBasics), University of Freiburg, Freiburg, Germany; Department of Psychosomatic Medicine and Psychotherapy, Medical Center, University of Freiburg; kbo-Inn-Salzach-Klinikum, Clinical Center for Psychiatry, Psychotherapy, Psychosomatic Medicine, Geriatrics and Neurology, Gabersee, Wasserburg/Inn, Germany; Department of Psychiatry and Psychotherapy III, University of Ulm, Ulm, Germany; German Center for Mental Health (DZPG), Site Mannheim-Heidelberg-Ulm, Germany; Department of Psychiatry and Psychotherapy, LVR-Klinikum Düsseldorf, Heinrich-Heine-Universität Düsseldorf, Medical Faculty, Düsseldorf, Germany; Department of Psychiatry and Psychotherapy, Charité-Campus Benjamin Franklin, Berlin, Germany; German Center for Mental Health (DZPG), Site Berlin-Potsdam, Germany; Department of Psychiatry, University of São Paulo Medical School, São Paulo, Brazil

**Author notes:** <u>Corresponding author:</u> Dr. Gerrit Burkhardt, Department of Psychiatry and Psychotherapy, LMU University Hospital, Nußbaumstraße 7, 80336 Munich, Germany, E-Mail. These authors equally contributed to the manuscript (Andre R Brunoni, Frank Padberg).

## Abstract

Machine-learning (ML) classification may offer a promising approach for treatment response prediction in patients with major depressive disorder (MDD) undergoing non-invasive brain stimulation. This analysis aims to develop and validate such classification models based on easily attainable sociodemographic and clinical information across two randomized controlled trials on transcranial direct-current stimulation (tDCS) in MDD. Using data from 246 patients with MDD from the randomized-controlled DepressionDC and ELECT-TDCS trials, we employed an ensemble machine learning strategy to predict treatment response to either active tDCS or sham tDCS/placebo, defined as ≥ 50% reduction in the Montgomery-Åsberg Depression Rating Scale at 6 weeks. Separate models for active tDCS and sham/placebo were developed in each trial and evaluated for external validity across trials and for treatment specificity across modalities. Additionally, models with above-chance detection rates were associated with long-term outcomes to assess their clinical validity. In the DepressionDC trial, models achieved a balanced accuracy of 63.5% for active tDCS and 62.5% for sham tDCS in predicting treatment responders. The tDCS model significantly predicted MADRS scores at the 18-week follow-up visit (F_(1,60)_ = 4.53, p_FDR_ = .037, R^2^ = 0.069). Baseline self-rated depression was consistently ranked as the most informative feature. However, response prediction in the ELECT-TDCS trial and across trials was not successful. Our findings indicate that ML-based models have the potential to identify responders to active and sham tDCS treatments in patients with MDD. However, to establish their clinical utility, they require further refinement and external validation in larger samples and with more features.

## Introduction

Major Depressive Disorder (MDD) represents a significant global health challenge, ranking as one of the main causes of disability worldwide^1^. Despite the availability of effective treatments ranging from pharmacotherapy and psychotherapy to non-invasive and invasive neurostimulation, many patients do not achieve remission, even after multiple therapeutic attempts^2^. The development of new interventions has proven challenging, possibly due to the heterogeneity of MDD symptoms^3^, its varying time course^4^, and a lack of robust biological correlates^5,6^. While multiple sociodemographic, clinical, genetic, and neuroimaging variables have been associated with responses to common treatments like antidepressant medication^7^, these associations have not yet resulted in stratified patient selection algorithms or targeted interventions. Thus, recent research has focused on developing multivariate predictive models that might enable pre-treatment stratification at the individual patient level^8,9^ and detect effects beyond the between-group level in randomized controlled trials (RCT)^9,10^. Within this approach, initial machine learning (ML)-based predictive models are typically trained on data from existing RCTs to identify responders to the treatments under investigation^8,11,12^. Models then require testing in independent samples and across diverse populations to ensure their generalizability to unseen patients before they are finally tested prospectively for clinical utility. However, efforts to externally validate initial models remain sparse^13^, and consequently, few attempts have been made to validate treatment prediction models in RCTs^14^.

Predictive approaches are particularly relevant in the field of non-invasive brain stimulation (NIBS), including repetitive transcranial magnetic stimulation (rTMS) and transcranial direct current stimulation (tDCS), as the clinical application of these interventions is rapidly growing. tDCS is a safe, well-tolerated, and easily applicable treatment option for patients with MDD^15–17^, but has yielded inconclusive results in recent confirmatory multicenter RCTs^18–20^. Therefore, efforts to optimize outcomes on the individual patient level are required to develop the intervention toward clinical applicability^21,22^. A recent study reported a high predictive accuracy of an ML-based prediction model for identifying responders to bifrontal transcranial direct current stimulation (tDCS), yet lacked external validation^23^. For rTMS, ML studies have mainly focused on other neuropsychiatric conditions, e.g. schizophrenia^9,10^. To our knowledge, there are currently no studies available utilizing ML models across RCTs on NIBS interventions.

To investigate whether sociodemographic and clinical data are informative for predicting the individual response to tDCS, we used data from two large RCTs on basically identical tDCS protocols (i.e. bifrontal electrode montage: anode left and cathode right dorsolateral prefrontal cortex [DLPFC], 2 mA intensity and 30 min duration), that were performed in Brazil^16^ and Germany^18^, evaluating the efficacy of tDCS in patients with MDD. We aimed to develop and externally validate ML-based prediction models to identify patients likely to benefit from tDCS, test those models for treatment specificity, and explore their clinical validity and utility based on long-term outcomes.

## Subjects and Methods

### Study design

In this secondary analysis of two randomized, blinded, sham-controlled trials, we used an ensemble ML-based strategy with nested cross-validation to identify patients with response to either active tDCS or sham/placebo treatment using sociodemographic and clinical baseline variables. Models were trained separately in each trial for each treatment modality and then applied 1.) across trials to test for external validity and 2.) across treatment modalities to test if predictions were specific to the treatment (see Figure 1). Classification probabilities of models with above-chance detection rates were then associated with long-term outcomes at follow-up to explore the clinical validity and utility of the predictions.

**Figure 1.**
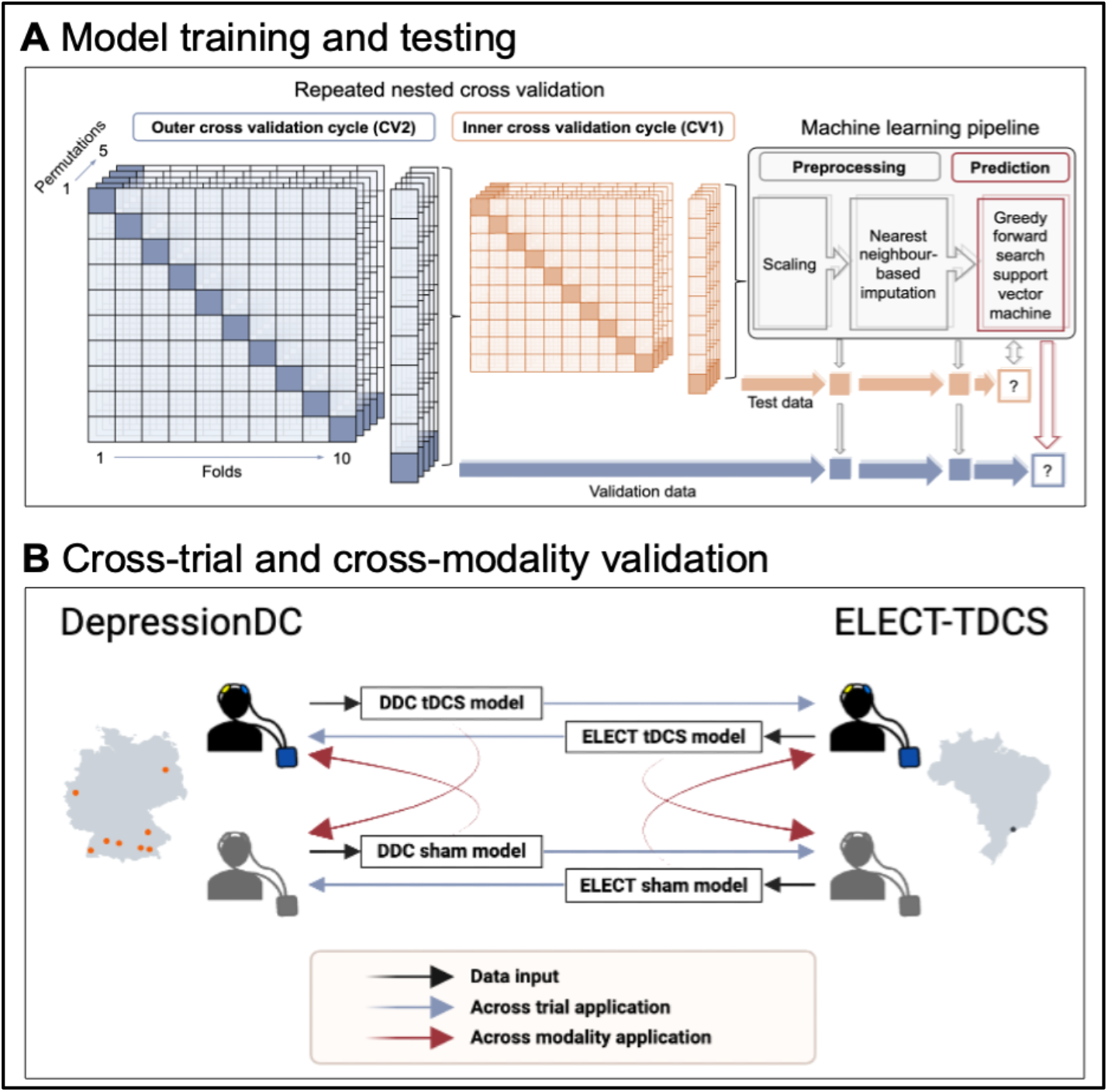
Model development and validation

### Study population

We analyzed patients with MDD from two trials: 1.) DepressionDC (trial registration: NCT02530164) was a multicenter RCT investigating the efficacy of 6 weeks of bifrontal tDCS as an additional treatment to selective serotonin reuptake inhibitors (SSRI) in patients with MDD^18^. Between January 2016 and June 2020, 160 patients were recruited at seven university hospitals and one psychiatric community hospital in Germany. Active tDCS was not superior to sham tDCS in reducing depressive symptoms. 2.) The Escitalopram versus Electrical Current Therapy for Treating Depression Clinical Study (ELECT-TDCS; trial registration: NCT01894815) was a single-center, non-inferiority RCT comparing active tDCS plus placebo medication, escitalopram plus sham tDCS, and sham tDCS plus placebo medication in patients with MDD over 10 weeks^16^. Two hundred forty-five patients were recruited at the University of São Paulo, Brazil, between October 2013 through July 2016. Active tDCS was not non-inferior to escitalopram but superior to placebo treatment.

Both trials employed rigorous RCT methods with stringent randomization, blinding protocols, and non-active sham conditions. Participants were selected based on the DSM-5 criteria for MDD while excluding patients with bipolar disorder, substance abuse or dependence, dementia, and personality disorders. However, DepressionDC enrolled patients currently receiving SSRIs, whereas ELECT-TDCS required participants to be antidepressant-free. Furthermore, DepressionDC permitted the inclusion of patients with marginally lower symptom severity, as assessed by the Hamilton Depression Rating Scale (HDRS), and restricted participation to a narrower age range. A comprehensive list of eligibility criteria for both trials is provided in the supplement.

The intervention followed a basically identical tDCS protocol with 2 mA stimulation of the DLPFC over 30 minutes per session. However, protocols differed in precise electrode placement, i.e. DepressionDC using the F3 (anode) and F4 (cathode) based on the international electroencephalogram 10-20 system, and ELECT-TDCS the Omni-Lateral-Electrode (OLE) system (left DLPFC anode and right DLPFC cathode) with slightly more lateral positions of electrodes^24^. Also, treatment lasted 10 weeks with a total of 22 treatment sessions (15 sessions in the first 3 weeks and 7 weekly sessions for the remaining treatment period) in ELECT-TDCS versus 6 weeks with a total of 24 treatment sessions (20 sessions in the first 4 weeks and 2 sessions per week for 2 weeks) in DepressionDC, with efficacy assessed at these time points using the HDRS and Montgomery-Åsberg Depression Rating Scale (MADRS) as primary outcome measures, respectively. Weekly MADRS scores were additionally collected in ELECT-TDCS as a secondary outcome measure.

We used data from all patients with available depression scores on the MADRS at baseline and week 6 after randomization. Thus, the analysis included 136 (active tDCS: 72 patients; sham tDCS: 64 patients) out of 150 patients from the DepressionDC sample and 110 (active tDCS + placebo: 66 patients; sham tDCS + placebo: 44 patients) out of 154 patients from the respective treatment arms of the ELECT-TDCS sample. All participants had provided their written informed consent before inclusion in the respective study. Both studies were approved by the local ethics committees and conducted in accordance with the Declaration of Helsinki.

### Prediction target and features

Participants were classified as treatment responders if they achieved a ≥ 50% reduction from baseline to week 6 on the 10-item MADRS (score range 0-60; higher scores indicate more severe depression)^25^. Pursuing a data-driven approach, we included all variables available across the studies at baseline as potential predictors. This amounted to 15 features, including basic sociodemographic information (age, sex, years of education, marriage, unemployment), medical history (body mass index, smoker status, diagnoses of hypertension, diabetes, and/or hypothyroidism), psychiatric history (age of MDD onset, duration of MDD episode, family history of MDD), and baseline depression severity (MADRS and Beck Depression Inventory-II [BDI-II] total scores).

### Machine learning analysis

All ML analyses were conducted using the in-house, open-source software package NeuroMiner, version 1.05 (https://github.com/neurominer-git/NeuroMiner-1), running on MATLAB (version R2022a). We used repeated nested cross-validation (CV) with 10 folds and 5 repetitions at both the inner (CV_1_) and outer (CV_2_) loops to strictly separate the training and testing of the models. In each CV_1_ fold, we scaled all features from 0 to 1 and substituted missing values via 7-nearest neighbor-based imputation (Euclidean distance for continuous, Hamming distance for categorical variables)^26^. Following a previous approach^27,28^, each processed CV_1_ training sample then entered a greedy stepwise forward search wrapper employing a linear support vector machine algorithm (SVM; LIBSVM 3.12^29^) to iteratively select a subset of 50% features with highest predictive performance (balanced accuracy 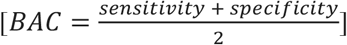 ^30^ on the held-out CV_1_ data) across a range of C hyperparameters 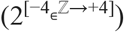. To account for uneven distributions of the outcome labels (response/non-response), optimal C hyperparameters were multiplied with the inverse ratio of the training group sizes^31^. For each CV_1_ permutation, all CV_1_ models were retrained with the optimal model hyperparameters, and this ensemble was then applied to the CV_2_ test data without modification. Classification probabilities for each CV_2_ test subject were retrieved by combining the decisions across all models. We calculated permutation-based *p*-values to define which models reached above-chance detection rates (α=0.05; 1000 permutations)^32^. To understand which features were most reliably contributing to the prediction of treatment response, we computed the CV ratio^33^. The feature importance of variables was further estimated using sign-based consistency mapping^34^. For cross-trial and cross-treatment modality validation, we applied CV_1_ ensembles with permutation-based above-chance detection rates without modification to the respective other samples. Out-of-sample performance metrics were calculated by comparing the predicted versus the observed outcome labels over all CV_2_ predictions.

### Post-hoc clinical validation

Additional validation analyses and visualizations were performed in R, version 4.3.2^35^. Results were considered significant at *α*=0.05. To explore the clinical validity of all classifiers with above-chance detection rates, we fit linear mixed models (LMM) using the lme4 package^36^ to predict MADRS and GAF changes from baseline until the trials’ follow-up appointments based on the models’ assigned probability to be a responder (formula: change ∼ assigned probability). The model included the treatment site as a random effect (formula: ∼1| site). The significance of the model factors was determined using omnibus tests (type III ANOVA) with Satterthwaite approximation to degrees of freedom.

## Results

### Main classifiers

In the DepressionDC trial, 24 patients (33%) had responded to active tDCS treatment at week 6. Compared to tDCS non-responders, these patients were significantly older (mean [SD] age: 37 [13] vs. 30 [13]; p=0.023) and showed lower clinician-rated (mean [SD] MADRS scores: 22 [5] vs. 26 [6]; p=0.049) and self-reported depression severity (mean [SD] BDI scores: 23 [9] vs. 30 [11]; p=0.011) at baseline (other baseline characteristics are shown in Table 1). The classifier ensemble predicted tDCS responders with an above-chance cross-validated BAC of 63.5% (P=0.001; Table 2; Figure 2), increasing the prognostic certainty compared to the base rate (prognostic summary index of 24.1%).

**Table 1.** Baseline characteristics of patients with MADRS response and non-response to tDCS

| Feature | DepressionDC |  |  | ELECT |  |  |
| --- | --- | --- | --- | --- | --- | --- |
|  | Non-responder,<br>n = 48<sup>a</sup> | Responder,<br>n = 24<sup>a</sup> | p-value<sup>b</sup> | Non-responders,<br>n = 39<sup>a</sup> | Responder,<br>n = 27<sup>a</sup> | p-value<sup>b</sup> |
| Sex |  |  | 0.9 |  |  | 0.3 |
| Female | 29 (60%) | 14 (58%) |  | 25 (66%) | 21 (78%) |  |
| Male | 19 (40%) | 10 (42%) |  | 13 (34%) | 6 (22%) |  |
| Age at randomization<br>- years | 37 (13) | 43 (14) | 0.1 | 44 (13) | 47 (11) | 0.2 |
| Age of depression<br>onset - years | 30 (13) | 37 (13) | **0.023** | 27 (12) | 28 (12) | 0.8 |
| Duration of episode -<br>weeks | 56 (67) | 52 (56) | >0.9 | 28 (72) | 26 (33) | 0.7 |
| Family history of<br>depression | 21 (46%) | 15 (65%) | 0.1 | 26 (67%) | 19 (70%) | 0.8 |
| Years of education | 11.91 (2.28) | 11.50 (1.65) | 0.5 | 15.8 (4.5) | 14.0 (4.7) | 0.2 |
| Unemployed | 3 (9.1%) | 0 (0%) | 0.5 | 13 (35%) | 10 (37%) | 0.9 |
| Married | 5 (15%) | 6 (43%) | 0.1 | 19 (49%) | 16 (59%) | 0.4 |
| Body mass index -<br>kg/m<sup>2</sup> | 25.7 (5.2) | 27.9 (5.9) | 0.1 | 25.8 (4.7) | 25.7 (4.6) | >0.9 |
| Smoker | 21 (44%) | 7 (29%) | 0.2 | 7 (18%) | 5 (19%) | >0.9 |
| Hypertension | 4 (10%) | 4 (24%) | 0.2 | 9 (24%) | 5 (19%) | 0.7 |
| Diabetes | 1 (2.5%) | 0 (0%) | >0.9 | 3 (7.9%) | 1 (3.7%) | 0.6 |
| Hypothyroidism | 2 (5.0%) | 4 (24%) | 0.1 | 8 (21%) | 5 (19%) | 0.8 |
| MADRS score at<br>baseline | 26 (6) | 22 (5) | **0.049** | 26 (6) | 29 (8) | **0.020** |
| BDI score at baseline | 30 (11) | 23 (9) | **0.011** | 30 (10) | 30 (11) | 0.7 |

<sup>a</sup>Mean (SD); n (%). <sup>b</sup> Pearson's Chi-squared test; Wilcoxon rank sum test. Fisher's exact test.

**Table 2.**
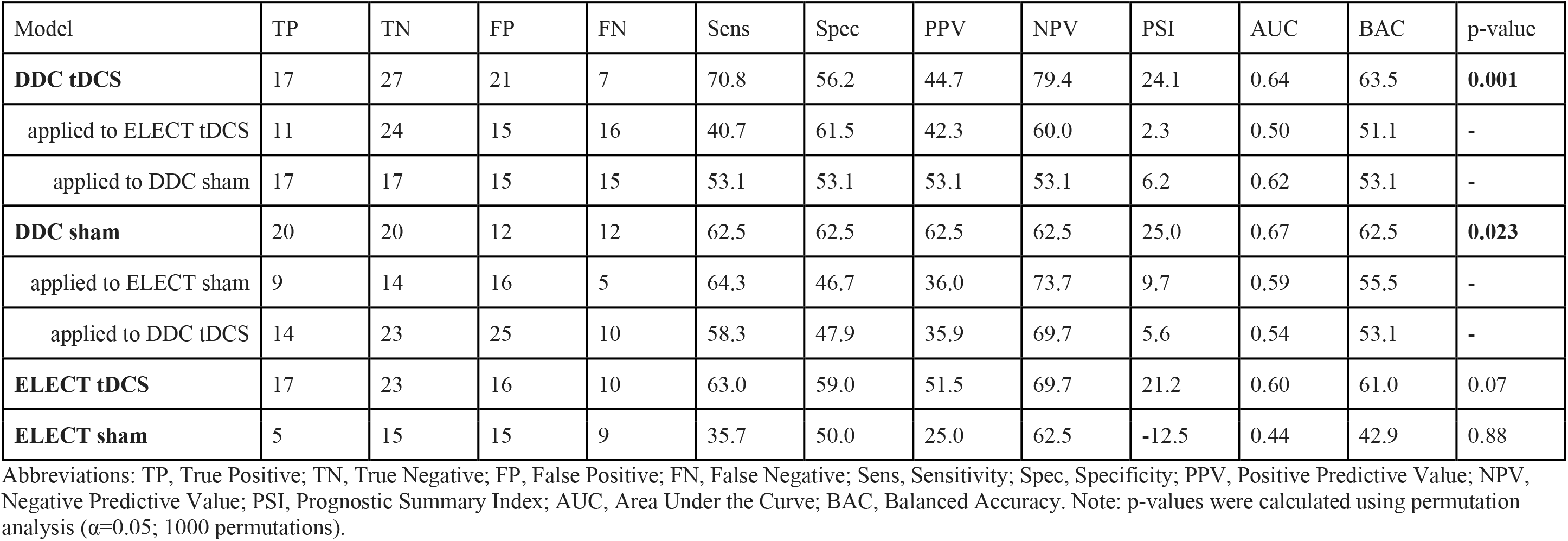
Prediction results of MADRS response models.

| Model | TP | TN | FP | FN | Sens | Spec | PPV | NPV | PSI | AUC | BAC | p-value |
| --- | --- | --- | --- | --- | --- | --- | --- | --- | --- | --- | --- | --- |
| **DDC tDCS** | 17 | 27 | 21 | 7 | 70.8 | 56.2 | 44.7 | 79.4 | 24.1 | 0.64 | 63.5 | **0.001** |
| applied to ELECT tDCS | 11 | 24 | 15 | 16 | 40.7 | 61.5 | 42.3 | 60.0 | 2.3 | 0.50 | 51.1 | - |
| applied to DDC sham | 17 | 17 | 15 | 15 | 53.1 | 53.1 | 53.1 | 53.1 | 6.2 | 0.62 | 53.1 | - |
| **DDC sham** | 20 | 20 | 12 | 12 | 62.5 | 62.5 | 62.5 | 62.5 | 25.0 | 0.67 | 62.5 | **0.023** |
| applied to ELECT sham | 9 | 14 | 16 | 5 | 64.3 | 46.7 | 36.0 | 73.7 | 9.7 | 0.59 | 55.5 | - |
| applied to DDC tDCS | 14 | 23 | 25 | 10 | 58.3 | 47.9 | 35.9 | 69.7 | 5.6 | 0.54 | 53.1 | - |
| **ELECT tDCS** | 17 | 23 | 16 | 10 | 63.0 | 59.0 | 51.5 | 69.7 | 21.2 | 0.60 | 61.0 | 0.07 |
| **ELECT sham** | 5 | 15 | 15 | 9 | 35.7 | 50.0 | 25.0 | 62.5 | -12.5 | 0.44 | 42.9 | 0.88 |

Abbreviations: TP, True Positive; TN, True Negative; FP, False Positive; FN, False Negative; Sens, Sensitivity; Spec, Specificity; PPV, Positive Predictive Value; NPV, Negative Predictive Value; PSI, Prognostic Summary Index; AUC, Area Under the Curve; BAC, Balanced Accuracy. Note: p-values were calculated using permutation analysis ( $\alpha=0.05$ ; 1000 permutations).

**Figure 2.**
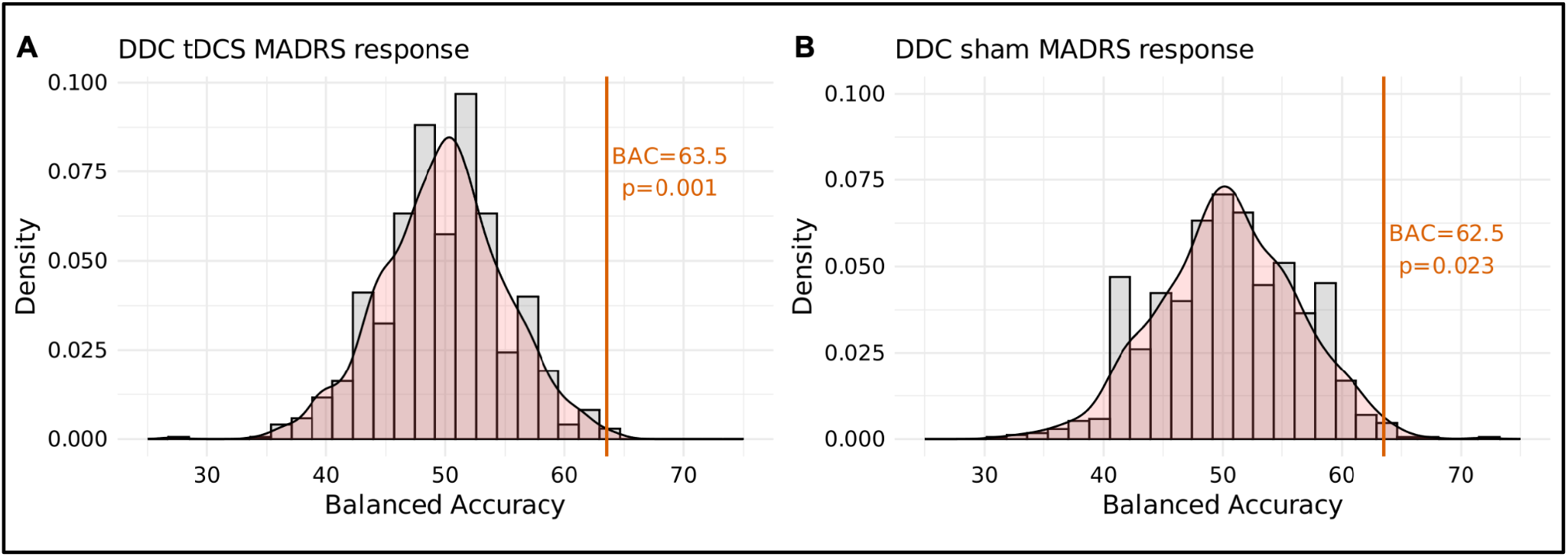
Permutation-based significance. Response was defined as ≥ 50% reduction from baseline.

In the sham group, 32 patients (50%) showed a treatment response. Compared to sham non-responders, these patients had lower clinician-rated (mean [SD] MADRS scores: 22 [5.3] vs. 24.3 [4.4]; p=0.017) and self-reported depression severity (mean [SD] BDI scores: 23 [9] vs. 30 [10]; p=0.005) at baseline. Sham responders were predicted with an above-chance cross-validated BAC of 62.5% (p=0.023; Table 2) and prognostic summary index of 25.1%. In both the tDCS and sham analyses, only baseline BDI scores reliably and significantly contributed to the classifier decisions (Figure 3).

**Figure 3.**
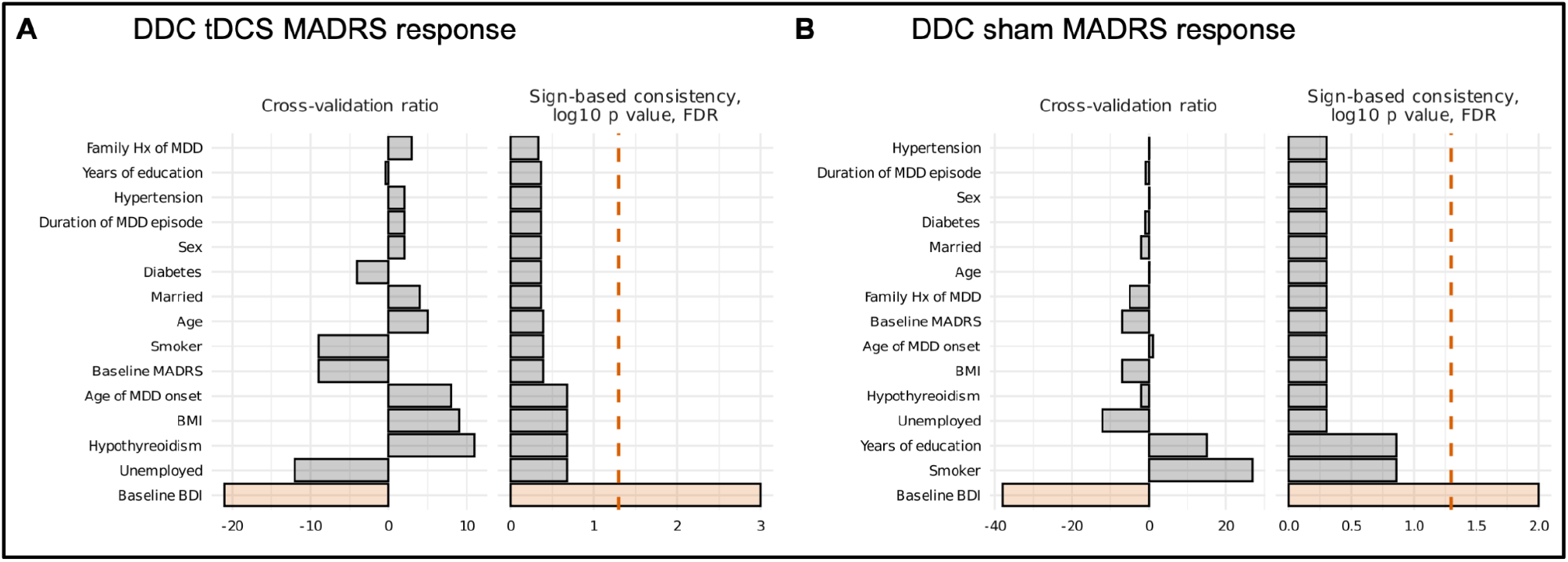
Feature importance. A positive cross-validation ratio suggests that a higher feature value predicts MADRS response, whereas a negative ratio implies that lower values do. A higher sign-based consistency suggests that feature weights were more consistently positive or negative across the ensemble. Significance was assessed by defining a hypothesis test for the importance score with a null hypothesis of 0 and an alternative hypothesis of not 0. FDR-corrected p-values were then calculated using a cumulative distribution function of z - scores (α=0.05; red dotted line).

In the ELECT trial, 27 patients (41%) had responded to active tDCS treatment at week 6. Compared to tDCS non-responders, they had higher clinician-rated depression at baseline (mean [SD] MADRS scores: 29 [8] vs. 26 [6]; p=0.020). Responders were predicted with a BAC of 61.0% that did not reach our above-chance detection criterion (p=0.071; Table 2). Similarly, our analysis did not yield models with above-chance detection rates for the 14 (32%) responders to sham treatment (BAC=42.9%; p=0.88; Table 2).

### External validation and treatment specificity

Models with above-chance detection rates did not generalize across trials. The DepressionDC tDCS classifier showed a BAC of 51.1% in the ELECT tDCS sample, and the DepressionDC sham classifier reached a BAC of 55.5% in the ELECT sham sample (shown in Table 2). Models also did not reach above-chance detection rates across treatment modalities, with the DepressionDC tDCS model showing a BAC of 53.1% when applied to the DepressionDC sham arm and the DepressionDC sham model showing a BAC of 53.1% when applied to the DepressionDC tDCS arm.

### Post-hoc clinical validation

For the DepressionDC tDCS model, classification probabilities significantly predicted MADRS scores at the 18-week follow-up visit (F_(1,60)_ = 4.53, p_FDR_ = .037, R^2^ = 0.069), but not MADRS scores at week 30 or GAF scores at weeks 18 and 30 (Supplementary Table S2 and Supplementary Figure S1). Classification probabilities of the DepressionDC sham model did not predict MADRS or GAF outcomes at weeks 18 and 30.

## Discussion

To our knowledge, this is the first study testing the cross-trial validity of ML models trained on easily attainable sociodemographic and clinical baseline variables to predict responders to a NIBS intervention. Whereas ML models increased accuracy in identifying responders to active tDCS and sham tDCS in the 6-week multicenter, randomized-controlled DepressionDC trial, the same variable battery could not be utilized to predict responses in the ELECT-TDCS trial, and models did not generalize across trial datasets and treatment modalities. Our findings underscore existing challenges and limitations inherent in predicting antidepressant responses in RCT populations.

The predictive accuracies of our models with above-chance detection rates align with prior attempts to predict responses to antidepressant medication using ML algorithms trained on clinical variables^9,11,12,37^. Although these performances are modest compared to the classification benchmarks set in other medical disciplines, such as neuroradiology^38^, antidepressant response prediction remains a challenging task relying on subjective judgment, and thus, even small increases in predictive accuracy might theoretically inform clinical decisions. However, as exemplified by the only prior attempt to prospectively assess the clinical utility of an antidepressant response classifier, which failed to improve treatment outcomes when applied as a decision-making tool^14^, strong indicators are needed to justify further development of classifiers beyond the proof-of-concept stage. In the context of our study, the tDCS response classifier for the DepressionDC sample significantly predicted depression severity at the 18-week follow-up visit, suggesting potential clinical validity. Nonetheless, given the failed external validation and the need for enhanced performance, further refinement and testing of the model would be needed to establish its efficacy and reliability^39^.

Recent research, including a validation attempt across trials on antipsychotic medication for schizophrenia^40^, suggests that three main reasons might have contributed to our models’ failed transferability across trial datasets. First, trial populations might have been too heterogeneous, including patients at different disorder stages or with nuanced differences in psychopathology profiles not captured by the broad DSM-5 inclusion criteria. Indeed, participants in the DepressionDC trial showed numerically later depression onset and longer mean depression episode duration compared to ELECT-TDCS^16,18^. By contrast, baseline depression severity was comparable between trials. Second, compared to a previous ML prediction study in the ELECT-TDCS cohort^23^, our models showed considerably lower predictive performance in the same dataset. Since our analysis aimed to develop generalizable prediction models across two RCT cohorts, we took several methodological choices that may partly explain this difference. Instead of an XGBoost classifier, we used a validated ensemble learning strategy applying SVM algorithms within a nested cross-validation framework. This framework was chosen because it has been applied in several multisite analyses and optimized to generate generalizable models^9,27,28^. We also limited the input variables to features available in both datasets. Consequently, this meant omitting data modalities like neuropsychological test results, electrophysiological data, and imaging measurements, which might have been needed to specifically detect patterns of response in the active treatment arm. For example, a recent analysis on data from the RESIS trial, which like the DepressionDC trial was also negative regarding its primary and secondary outcomes, extended a previous attempt to built an active rTMS treatment response classifier for patients with predominant negative-symptom schizophrenia based on structural Magnetic Resonance Imaging (sMRI)^9^ by incorporating further data domains (i.e. polygenic risk scores) and multimodal sequential modelling^10^. While not yet validated, this approach improved the prediction performance from 80 % to 94% in the active treatment but not the sham treatment arm. Third, treatment response rates could have been overly influenced by contextual factors that cannot be modeled at the single-subject level. For example, the present trials were conducted in healthcare settings with differing models of reimbursement and access to care. They also subtly differed in eligibility criteria, with participants in the DepressionDC trial kept on a stable SSRI dose while participants in ELECT-TDCS were antidepressant-free. These methodological challenges showcase the current need in ML-based treatment prediction research to systematically assess and compare potential analytic pipelines in larger samples, to include comprehensive phenotyping in RCT protocols, and to harmonize best-practice symptom assessments across brain stimulation trials.

Models with above-chance detection rates for active tDCS and sham tDCS in the DepressionDC sample also did not generalize across treatment modalities. Our feature importance analyses indicated that the performance of both models was predominantly driven by baseline BDI scores. This reliance on a single feature presents an interpretative challenge: Without a distinct feature selection profile, it becomes difficult to determine whether the models are tailored specifically to each treatment modality or if they lack generalizability to new patients.

Our analysis has several limitations. Firstly, the relatively small sample size in both datasets might have limited the performance and robustness of our classifiers^41^. Secondly, only a limited set of identical features was available from both trials. For example, negative affect, which was a key predictive feature in the prior ML analysis of the ELECT-TDCS sample^23^, was not collected in DepressionDC. This omission might have reduced the predictive accuracy of models in the present analysis. Thirdly, our study was retrospective and served as a proof-of-concept analysis.

Consequently, our models have not been validated prospectively, nor have they been benchmarked against clinical judgments. Fourthly, the DepressionDC trial did not demonstrate the efficacy of active tDCS at a group level. This raises the possibility that there may not have been any discernible effects at the individual subject level either, which would inherently limit the potential of our models to identify specific treatment effects. Lastly, while Kambeitz et al.^23^ evaluated treatment response at week 10, we opted to identify responders at week 6 due to the availability of MADRS data at this time point across both trials. Consequently, our study could not explore and compare predictive accuracies at various endpoints.

In conclusion, our findings suggest that readily accessible clinical variables at baseline, particularly self-rated depression severity, have the potential to identify responders to active tDCS and sham tDCS treatments in patients with MDD. However, our findings also caution against the premature dissemination of predictive models that lack external validation. Future research should aim to harmonize and deepen phenotyping efforts in RCT protocols to enable the development of more robust predictive models. Ultimately, such models need to be first externally validated and then prospectively tested for their clinical utility.

## Supporting information

Supplemental Methods and Results

## Data Availability

All data produced in the present study are available upon reasonable request to the authors.

## Acknowledgements

This research was funded by grant 01EE1403E within the German Center for Brain Stimulation research consortium by the German Federal Ministry of Education and Research and supported within the initial phase of the German Center for Mental Health (Deutsches Zentrum für Psychische Gesundheit [DZPG], grant 01EE2303A). GB was supported by two internal grants for young researchers from the Medical Faculty of LMU Munich (FöFoLe, grant number 1127; FöFoLe+, grant number CS063). JW was supported by an internal grant for young researchers from the Medical Faculty of LMU Munich (FöFoLe, grant number 1150).

