## Supplemental Methods and Results for "Cross-trial prediction of treatment response to transcranial direct current stimulation in patients with major depressive disorder"

### Supplementary methods and results

### Supplementary methods

**List of eligibility criteria for the DepressionDC trial**

Inclusion criteria:

- Men and women aged 18 to 65 years
- Primary DSM-5 diagnosis of major depression, as assessed by the Structured Clinical Interview for DSM-5 Axis I Disorders, Research Version (SCID-5-RV), with a single or recurrent episode and with the additional requirements of a current episode with a duration of at least 4 weeks
- Duration of current depressive episode is less than 5 years (the definition of an episode is demarcated by a period of ≥2 months in which the patient did not meet the full criteria for the DSM-5 definition of a major depressive episode)
- Total HDRS-21 score greater than or equal to 15 at the screening visit
- In the current episode, the patient did not respond to at least one antidepressant treatment, i.e. a minimum of 1 and a maximum of 4 antidepressant drug trials, of adequate dose and duration (defined as a minimum level of 2 on the Antidepressant Treatment History Form [ATHF])
- In the current episode, patient has been taking a selective serotonin reuptake inhibitor of adequate dose for at least 4 weeks (defined as a minimum level of 2 on the ATHF)
- Capable and willing to provide informed consent
- Negative pregnancy test and willingness to use contraceptive measures during study treatment for women of childbearing potential (i.e. <2 years after menopause)

Exclusion criteria:

- Investigator or site personnel directly affiliated with the study or immediate family member of such people (immediate family is defined as a spouse, parent, child, or sibling, whether by birth or legal adoption)
- Acute risk for suicide (Montgomery-Åsberg Depression Rating Scale [MADRS], item 10 score >4 or as assessed by the C-SSRS, agree with item 4 and/or item 5)
- High degree of treatment resistance, defined as >4 adequate treatment attempts in the current episode (each attempt with an ATHF score of >3)
- Treatment with electroconvulsive therapy in the present episode
- Treatment with deep brain or vagus nerve stimulation and/or any other intracranial implants (clips, cochlear implants)
- Any other relevant psychiatric axis I and/or axis II disorder, as assessed by the M.I.N.I. and the Structured Clinical Interview for DSM-IV Axis II Personality Disorders
- Any relevant unstable medical condition
- History of clinical tDCS treatment (except single experimental tDCS sessions)
- Pregnancy

**List of eligibility criteria for the ELECT trial**

Inclusion criteria:

- Men and women aged 18 to 75 years
- Unipolar depression (diagnosed according to DSM-5 criteria and confirmed by psychiatrists by means of the M.I.N.I.)
- Total HDRS-17 score greater than or equal to 17
- Low risk of suicide (evaluated with the use of the M.I.N.I.)

Exclusion criteria:

- bipolar disorder
- substance abuse or dependence
- dementia
- personality disorder
- brain injury
- pregnancy
- specific contraindications to tDCS (e.g., cranial plates)
- current or previous escitalopram use
- previous or concomitant participation in other trials of tDCS

### Supplementary results

**Supplementary Table S1: Baseline characteristics of patients with MADRS response and non-response to sham treatment**

| Feature | DepressionDC | | | ELECT | | |
|---|---|---|---|---|---|---|
| | Non-responder, n = 32[a] | Responder, n = 32[a] | p-value[b] | Non-responders, n = 30[a] | Responders, n = 14[a] | p-value[b] |
| Sex | | | 0.3 | | | 0.2 |
| Female | 20 (62%) | 16 (50%) | | 24 (80%) | 8 (57%) | |
| Male | 12 (38%) | 16 (50%) | | 6 (20%) | 6 (43%) | |
| Age at randomization - years | 40 (13) | 38 (14) | 0.5 | 41 (14) | 42 (13) | 0.6 |
| Age of depression onset - years | 33 (14) | 33 (14) | >0.9 | 24 (11) | 26 (12) | 0.6 |
| Duration of episode - weeks | 47 (51) | 48 (67) | 0.7 | 23 (25) | 34 (59) | >0.9 |
| Family history of depression | 17 (57%) | 16 (53%) | 0.8 | 21 (70%) | 11 (79%) | 0.7 |
| Years of education | 11.21 (1.67) | 11.79 (1.84) | 0.2 | 15.7 (3.8) | 16.7 (5.2) | 0.8 |
| Unemployed | 7 (35%) | 7 (35%) | 0.1 | 9 (31%) | 3 (23%) | 0.7 |
| Married | 3 (12%) | 1 (5.0%) | 0.6 | 9 (30%) | 4 (29%) | >0.9 |
| Body mass index - kg/m2 | 29 (9) | 26 (5) | 0.7 | 26.4 (5.9) | 26.1 (5.0) | 0.9 |
| Smoker | 8 (25%) | 14 (44%) | 0.1 | 6 (20%) | 1 (8.3%) | 0.7 |
| Hypertension | 3 (11%) | 2 (9.1%) | >0.9 | 8 (27%) | 2 (14%) | 0.5 |
| Diabetes | 1 (3.7%) | 1 (4.5%) | >0.9 | 1 (3.3%) | 1 (7.1%) | 0.5 |
| Hypothyreoidism | 3 (11%) | 1 (4.5%) | 0.6 | 1 (3.6%) | 0 (0%) | >0.9 |
| MADRS score at baseline | 24.3 (4.4) | 21.5 (5.3) | **0.017** | 28 (8) | 27 (5) | 0.4 |
| BDI score at baseline | 30 (10) | 23 (9) | **0.005** | 35 (12) | 27 (12) | 0.1 |

[a] Mean (SD); n (%). [b] Pearson's Chi-squared test; Wilcoxon rank sum test. Fisher's exact test.

**Supplementary Table S2: Prediction of follow-up outcomes.**

| DepressionDC tDCS model | Classification probability | | |
|---|---|---|---|
| | F (df[a]) | p[b] | $R^2$ |
| MADRS change at week 18 | 4.53 (1, 60) | **0.037** | 0.069 |
| MADRS change at week 30 | 2.19 (1, 54) | 0.1 | 0.036 |
| GAF change at week 18 | 0.02 (1, 53) | 0.9 | <0.001 |
| GAF change at week 30 | 0.03 (1, 49) | 0.9 | 0.001 |
| DepressionDC sham model | Classification probability | | |
| | F(df) | p | $R^2$ |
| MADRS change at week 18 | 0 (1,50) | 0.9 | <0.001 |
| MADRS change at week 30 | 1.68 | 0.2 | 0.033 |
| GAF change at week 18 | 0.35 (1, 48) | 0.6 | 0.348 |
| GAF change at week 30 | 0.11 (1, 47) | 0.8 | 0.002 |

[a] df=degrees of freedom. [b] p-value determined using type III ANOVA) with Satterthwaite approximation to degrees of freedom.

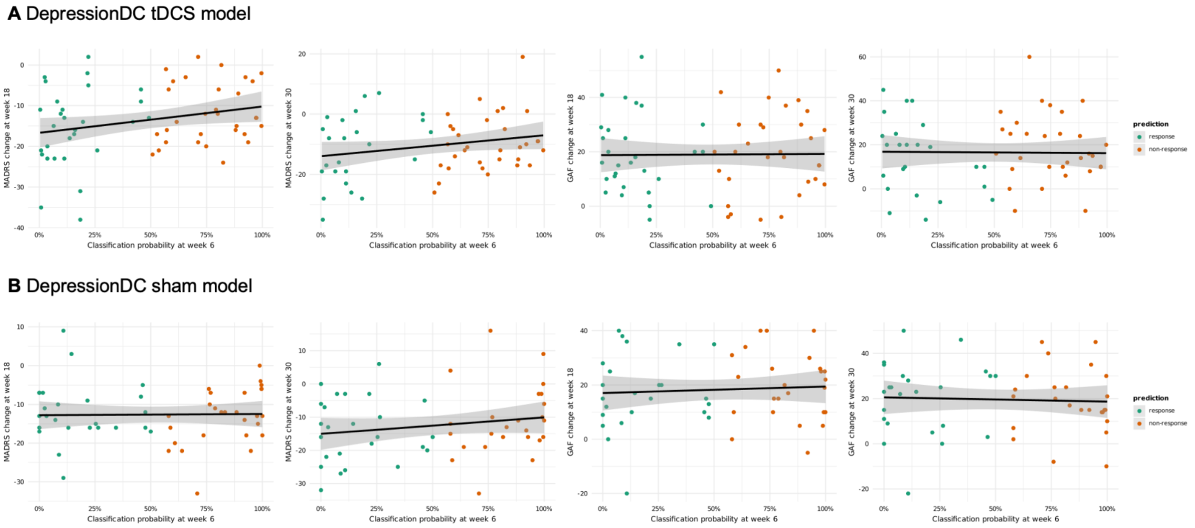

**Supplementary Figure S1:** Association between the classification probability of each individual and the follow-up scores on the MADRS and GAF at weeks 18 and 30 for the significant classification models.
